# SARS-CoV-2 Seroprevalence Across a Diverse Cohort of Healthcare Workers

**DOI:** 10.1101/2020.07.31.20163055

**Authors:** Joseph E. Ebinger, Gregory J. Botwin, Christine M. Albert, Mona Alotaibi, Moshe Arditi, Anders H. Berg, Aleksandra Binek, Patrick Botting, Justyna Fert-Bober, Jane C. Figueiredo, Jonathan D. Grein, Wohaib Hasan, Mir Henglin, Shehnaz K. Hussain, Mohit Jain, Sandy Joung, Michael Karin, Elizabeth H. Kim, Dalin Li, Yunxian Liu, Eric Luong, Dermot P.B. McGovern, Akil Merchant, Noah Merin, Peggy B. Miles, Margo Minissian, Trevor-Trung Nguyen, Koen Raedschelders, Mohamad A. Rashid, Celine E. Riera, Richard V. Riggs, Sonia Sharma, Sarah Sternbach, Nancy Sun, Warren G. Tourtellotte, Jennifer E. Van Eyk, Kimia Sobhani, Jonathan G. Braun, Susan Cheng

## Abstract

**Importance:** Antibody testing is important for understanding patterns of exposure and potential immunity to SARS-CoV-2. Prior data on seroprevalence have been subject to variations in selection of individuals and nature as well as timing of testing in relation to exposures.

**Objective:** We sought to determine the extent of SARS-CoV-2 seroprevalance and the factors associated with seroprevelance across a diverse cohort of healthcare workers.

**Design:** Observational cohort study of healthcare workers, including SARS-CoV-2 serology testing and participant questionaires.

**Participants:** A diverse and unselected population of adults (n=6,062) employed in a multi-site healthcare delivery system located in Los Angeles County, including individuals with direct patient contact and others with non-patient-oriented work functions.

**Exposure:** Exposure and infection with the SARS-CoV-2 virus, as determined by seropositivity.

**Main Outcomes:** Using Bayesian and multi-variate analyses, we estimated seroprevalence and factors associated with seropositivity and antibody titers, including pre-existing demographic and clinical characteristics; potential Covid-19 illness related exposures; and, symptoms consistent with Covid-19 infection.

**Results:** We observed a seroprevalence rate of 4.1%, with anosmia as the most prominently associated self-reported symptom in addition to fever, dry cough, anorexia, and myalgias. After adjusting for potential confounders, pre-existing medical conditions were not associated with antibody positivity. However, seroprevalence was associated with younger age, Hispanic ethnicity, and African-American race, as well as presence of either a personal or household member having a prior diagnosis of Covid-19. Importantly, African American race and Hispanic ethnicity were associated with antibody positivity even after adjusting for personal Covid-19 diagnosis status, suggesting the contribution of unmeasured structural or societally factors. Notably, number of people, or children, in the home was not associated with antibody positivity.

**Conclusion and Relevance:** The demographic factors associated with SARS-CoV-2 seroprevalence among our healthcare workers underscore the importance of exposure sources beyond the workplace. The size and diversity of our study population, combined with robust survey and modeling techniques, provide a vibrant picture of the demographic factors, exposures, and symptoms that can identify individuals with susceptibility as well as potential to mount an immune response to Covid-19.

**Key Points:** *Question:* What is the SARS-CoV-2 IgG seroprevalence rate across a large and diverse healthcare worker population, and which clinical, envionrmental, and symptom-based measures are associated with seropositivity?

*Findings:* We observed a seroprevalence rate of 4.1%. Adjusting for potential confounders, seropositivity was associated with younger age, Hispanic ethnicity, African-American race, and the symptom of anosmia, while not significantly associated with any pre-existing medical conditions.

*Meaning:* Factors associated with SARS-CoV-2 seroprevalence among our healthcare workers underscore the importance of exposure sources beyond the workplace.

## INTRODUCTION

Amidst the ongoing global pandemic caused by SARS-CoV-2, the viral agent causing Covid-19, substantial attention^1^ turned to antibody testing as an approach to understanding patterns of exposure and immunity across populations. The use and interpretion of antibody testing to assess exposure and immunity remains frought with inconsistencies and unclear clinical correlations, in part due to a dearth of high quality studies among diverse participants.^2,3^ Recent publications have pointed to the challenges and importance of understanding how different antibody tests for SARS-CoV-2 perform, and factors that may render one method superior to another.^4,5^ Nonetheless, there remains general agreement that antibody testing offers valuable information regarding the probable extent of SARS-CoV-2 exposure, the factors associated with exposure, and the potential nature and determinants of seropositive status.^6^

To the end, we conducted a study of SARS-CoV-2 antibody screening of a large, diverse, and unselected population of adults employed in a multi-site healthcare delivery system located in Los Angeles County, including individuals with direct patient contact and others with non-patient-oriented work functions. Recognizing the range of factors that might influence antibody status in a given individual, we focused our study on not only estimating seroprevalence but also on identifying factors associated with seropositivity and relative antibody levels within the following three categories: (1) pre-existing demographic and clinical characteristics; (2) potential Covid-19 illness related exposures; and, (3) Covid-19 illness related response variables (i.e. different types of self-reported symptoms).

## METHODS

### Study Sample

The sampling strategy for our study has been described previously.^7^ In brief, beginning on May 11, 2020, we enrolled a total of N=6,318 active employees working at multiple sites comprising the Cedars-Sinai Health System, located in the diverse metropolis of Los Angeles County, California. The Cedars-Sinai organization includes two hospitals (Cedars-Sinai Medical Center and Marina Del Rey Hospital) in addition to multiple clinics in the Cedars-Sinai Medical Delivery Network. All active employees (total N∼15,000) were invited to participate in the study by providing a peripheral venous blood sample for serology testing and completing an electronic survey of questions regarding past medical history, social history, and work environment in addition to Covid-19 related symptoms and exposures.^8,9^ For the current study, we included all participants who completed both SARS-CoV-2 antibody testing and electronic survey forms (N=6,062). The study protocol was approved by the Cedars-Sinai institutional review board and all participants provided written informed consent.

### Serologic Assays

All participant biospecimens underwent serology testing by the Cedars-Sinai Department of Pathology and Laboratory Medicine using the Abbott Diagnostics SARS-CoV-2 IgG chemiluminescent microparticle immunoassay assay (Abbott Diagnostics, Abbott Park, IL) performed on an Abbott Diagnostics Architect ci16200 analyzer. The assay reports a signal-to-cutoff ratio (S/CO) corresponding to the relative light units produced by the test sample compared to the relative light units produced by an assay calibrator sample. The manufacturer recommended S/CO ratio of 1.4 was used to assign binary seropositivity status. This cutoff was validated for high specificity (i.e., >99%) ∼14 days post symptom onset.^10^ The Abbott assay detects antibodies directed against the nucleocapsid (N) antigen of the SARS-CoV-2 virus, which assists with packaging the viral genome after replication, and achieves specificity for IgG by incorporating an anti-human IgG signal antibody.

### Statistical Analyses

#### Estimates of Seroprevalence

We conducted a literature review to identify published data (until June 25, 2020) on the sensitivity and specificity of the Abbott Architect SARS-CoV-2 IgG assay, applied in specific populations using the manufacturer’s recommended thresholds. We identified a total of 15 studies assessing sensitivity in 2,114 tests and 18 studies reporting specificity in 7,748 tests (**Supplemental Tables 1-2**); we combined this information with data from an additional independent cohort of 60 case and 178 control specimens used to asses sensitivity and specificity, respectively, within the Cedars-Sinai Department of Pathology and Laboratory Medicine. We noted that studies investigating specificity generally assessed samples collected prior to the SARS-CoV-2 pandemic whereas studies reporting sensitivity included specimens from RT-PCR confirmed individuals (see details provided in **Supplemental Tables 1-2**). We restricted our analyses to a referent cohort of tests conducted on samples from individuals who were assayed ≥7 days following symptoms onset to most closely match our cohort sample characteristics and the situational context for study enrollment. We integrated source population-level demographic data, representative of the entire Cedars-Sinai employee base, with data from our enrolled study sample using an Iterative Proportional Fitting procedure (IPF) to estimate the number of eligible employees within each demographic category (with provided population totals considered the target, using constraints derived from our sample).^11^ We then fit a Bayesian multilevel hierarchical logistic regression model using RStan,^12,13^ including reported age, gender, race/ethnicity and site as coefficients, to model exposure probability (see **Supplemental Methods** for full details). We estimated the seroprevalence within each post-stratified demographic category based on the averaged and weighted value of the expected number of employees within that category.

**Table 1.**
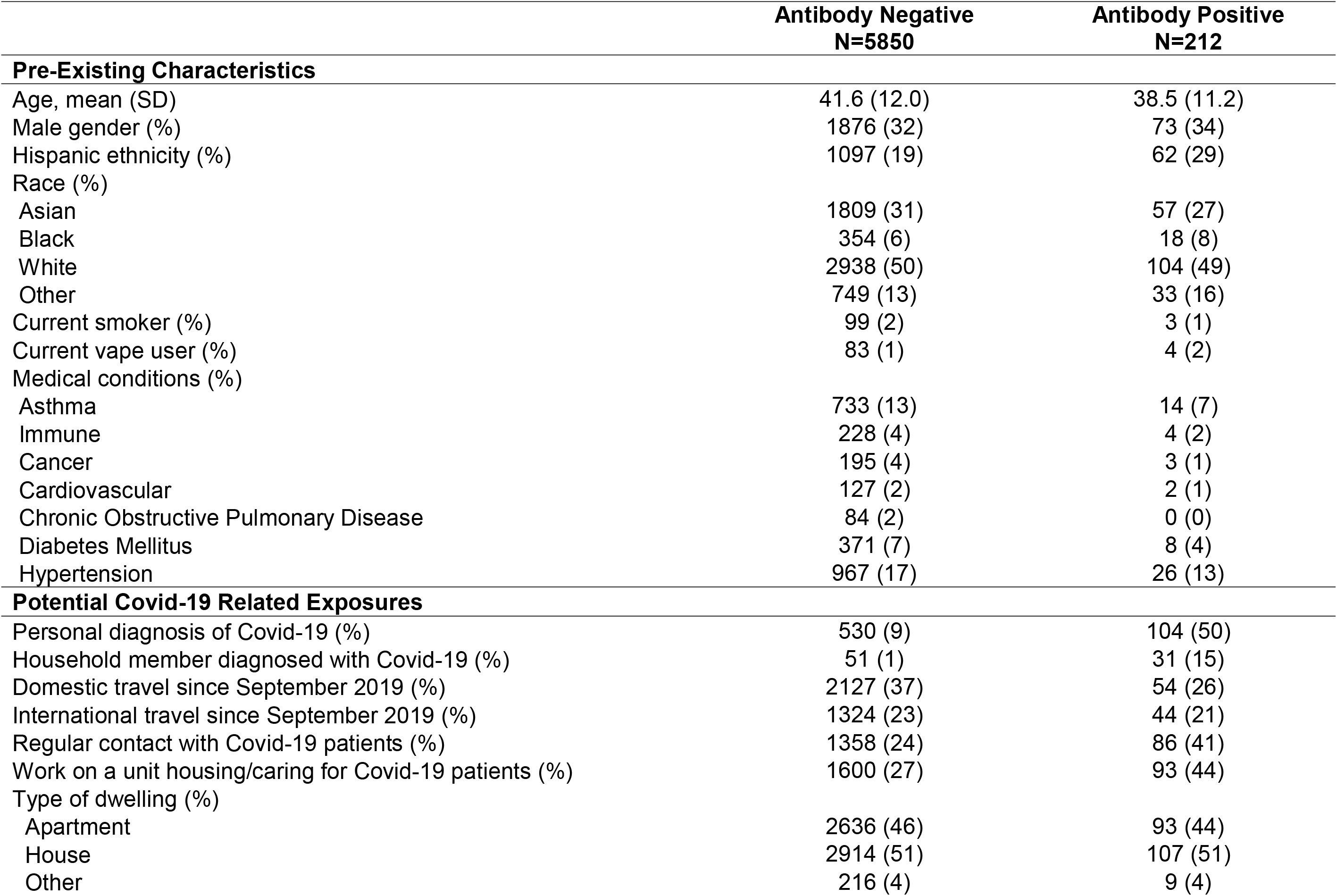
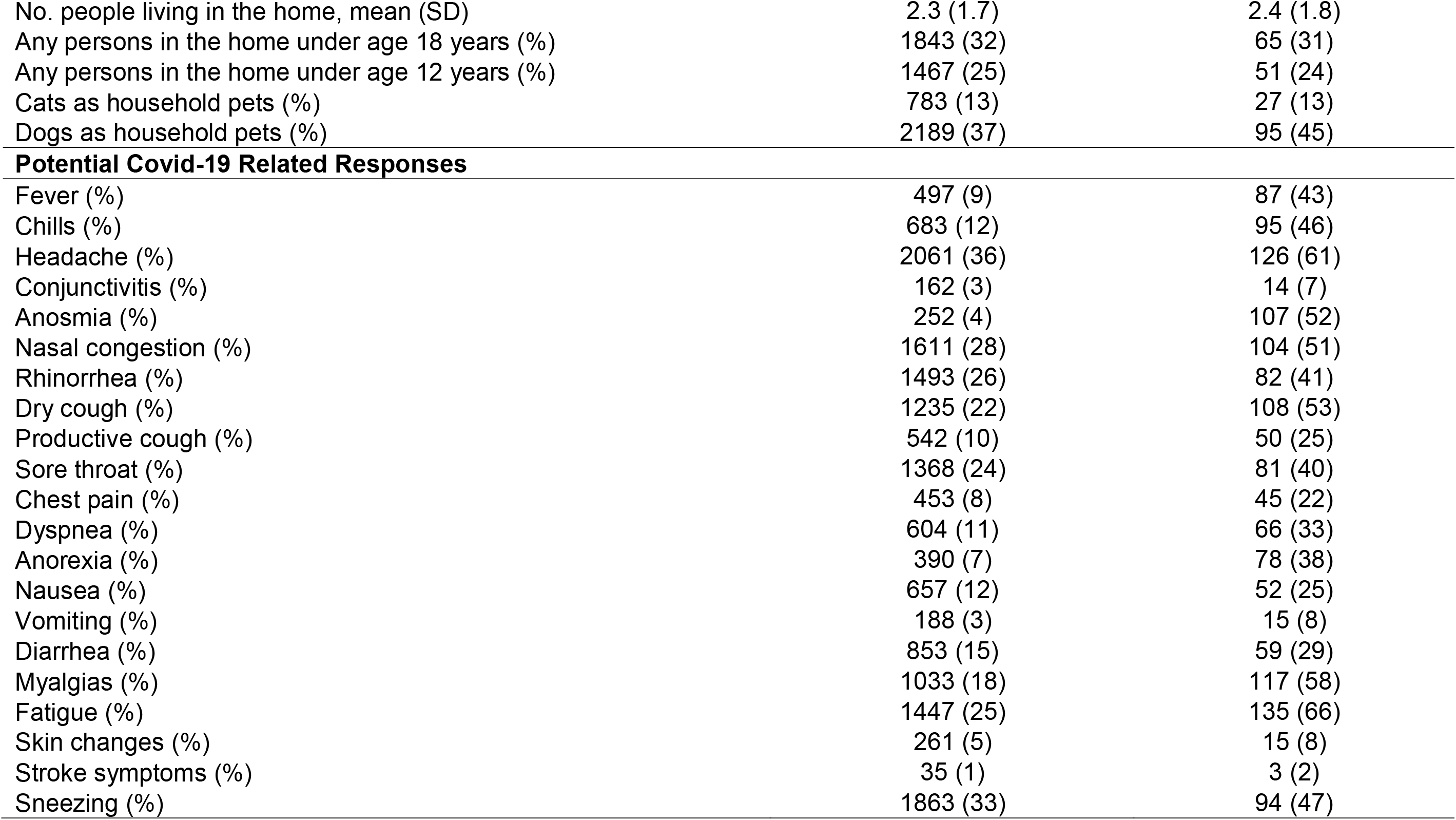
Characteristics of the Study Sample

#### Factors Associated with Seroprevalence

Prior to multivariable-adjusted analyses, age and IgG index were transformed by dividing by 10 for interpretability of coefficients in all models. In adjusted analyses, we compared differences between serology status (i.e. antibody positive versus negative) in each variable of interest, grouped into one of three categories: (1) pre-existing demographic and clinical characteristics (e.g. age, gender, ethnicity, race, and self-reported medical comorbidities); (2) Covid-19 related exposures (e.g. self-reported medical diagnosis of Covid-19 illness, household member with Covid-19 illness, number of people living in the home including children, type of home dwelling, etc); and, (3) Covid-19 related response variables (e.g. self-reported fever, chills, dry cough, anosmia, nausea, myalgias, etc.). In multivariable-adjusted analyses, we used logistic and linear models to examine the extent to which the three categories of variables (predictors) may be associated with antibody positive status (primary outcome) in the total sample or IgG antibody level in the subset of persons with positive antibody status (secondary outcome). Initial models were deliberately sparse, adjusting for a limited number of key covariates (e.g. age, gender) and those variables with associations meeting a significance threshold of P<0.10 were advanced for inclusion in a final multivariable model with only other variables identified from the sparse regression included. A final separate multivariable model was constructed for each of the 3 categories of variables.

## RESULTS

The demographic, clinical, exposure, and symptom response characteristics of the study sample are shown in **Table 1**, by antibody test result status; the study sample included individuals whose residence spanned diverse regions across Los Angeles County (**Supplemental Figure 1**). The overall seroprevalence was 4.1% (95% CI 3.1%, 5.7%), with higher estimates seen in younger compared to older individuals and in Hispanics compared to non-Hispanics (**Figure 1** and **Supplemental Table 3**).

**Figure 1.**
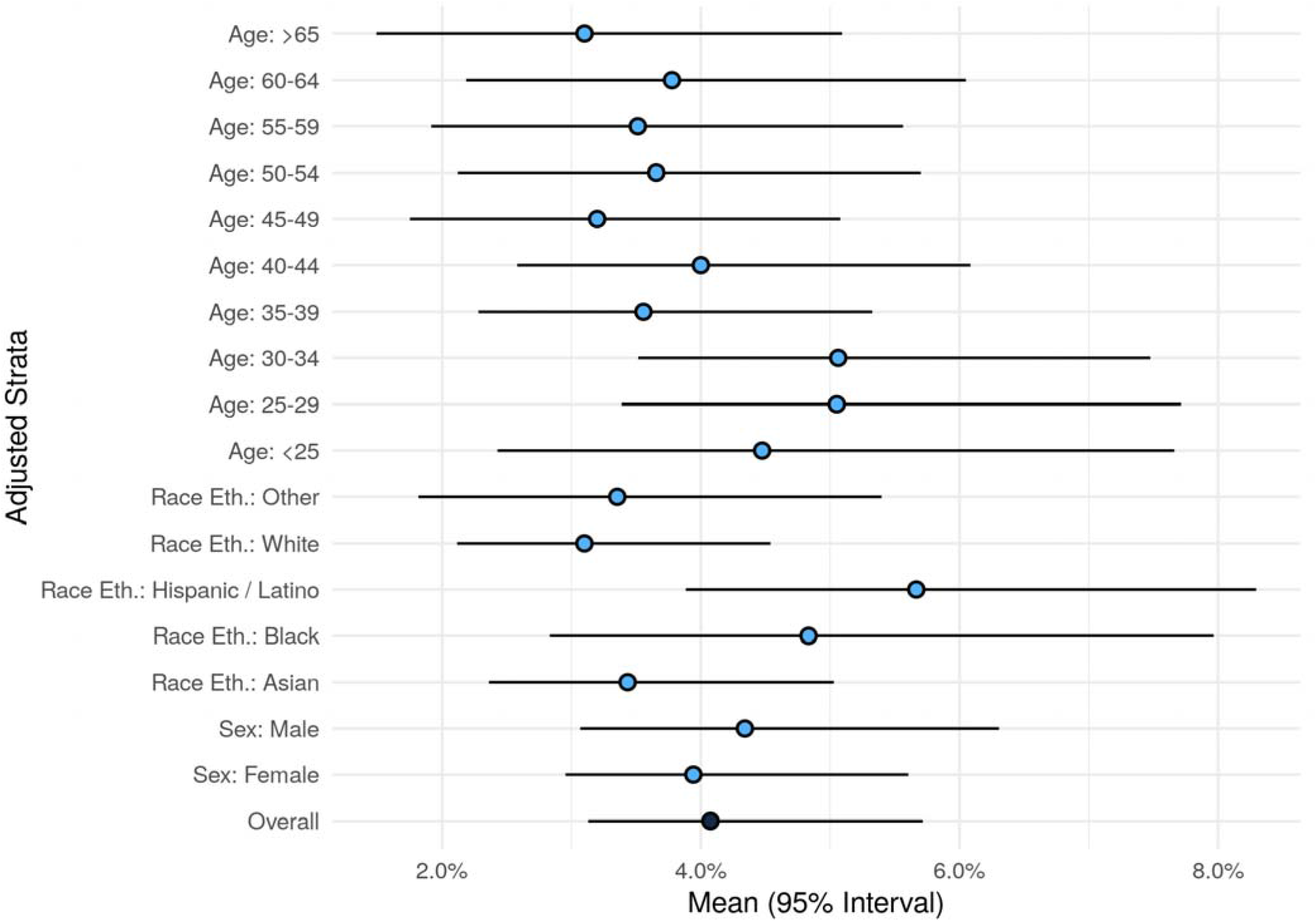
Seroprevalence Overall and by Subgroup.

In multivariable-adjusted analyses of pre-existing characteristics (**Figure 2** and **Supplemental Table 4**), the main factors significantly associated with greater odds of seropositive status were Hispanic ethnicity (OR 1.80 [95% CI 1.31, 2.46], P<0.001), and African American race (1.72 [1.03, 2.89], P=0.04), compared to non-Hispanic Whites. The main factors associated with lower odds of being seropositive were older age (0.81 [0.71, 0.92] per age decade, P=0.001), and a history of asthma (0.48 [0.26, 0.80], P=0.009). Among all seropositive persons, hypertension was significantly associated with higher antibody level (beta 0.11 [SE 0.04] per 10-unit increment in the IgG index, P=0.011).

**Figure 2.**
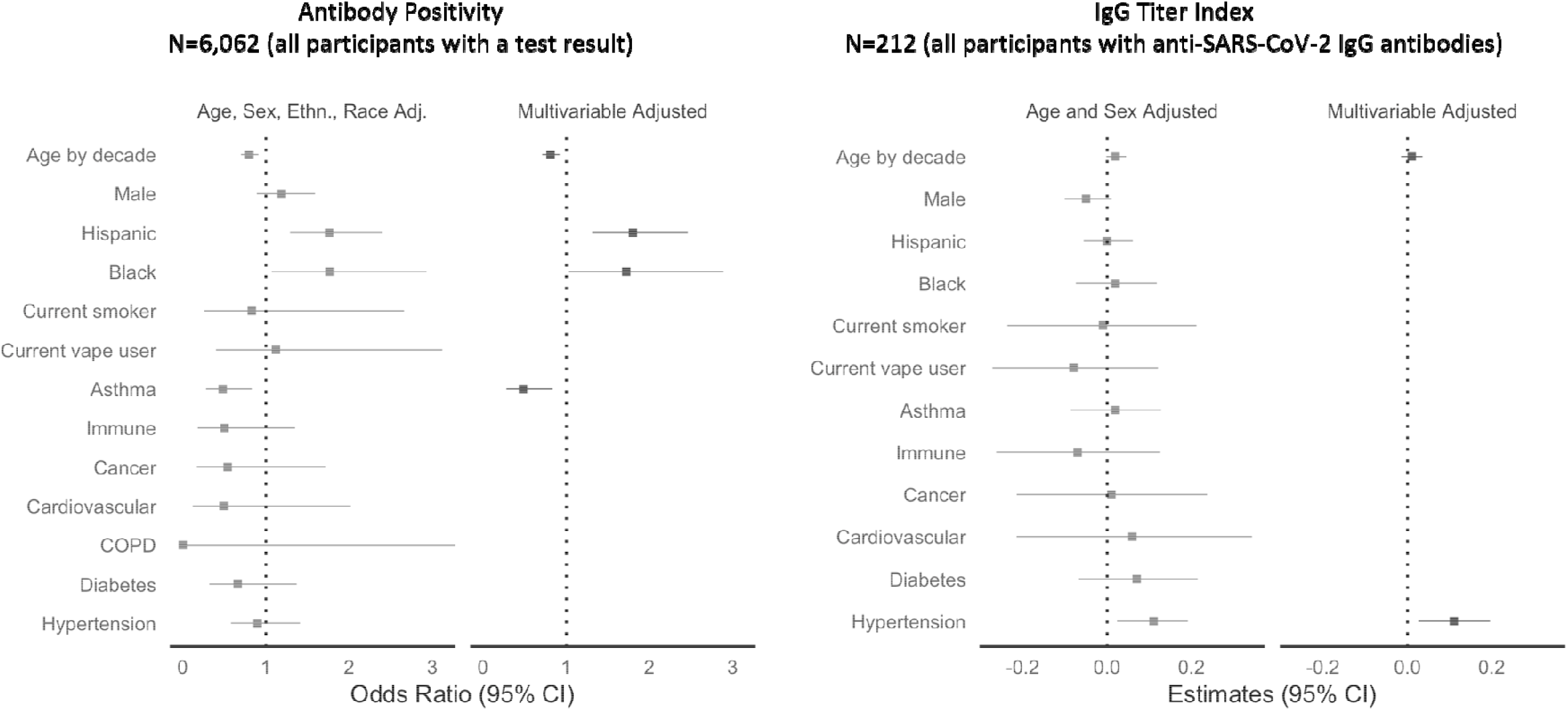
Pre-Existing Factors Associated with SARS-CoV-2 Seroprevalence.

In multivariable-adjusted analyses of Covid-19 related exposures (**Figure 3** and **Supplemental Table 5**), the factors significantly associated with greater odds of seropositive status were having had a medical diagnosis of Covid-19 (7.78 [5.73, 10.56], P<0.001) and a household member previously diagnosed with Covid-19 (9.42 [5.50, 16.13], P<0.001), with a similar trend observed for working in a location where Covid-19 patients are treated (1.61 [1.18, 2.18], P=0.002]. Among seropositive individuals, having a medical diagnosis of Covid-19 was associated with higher antibody level. Notably, dwelling type, number of people in the home, and having children or common domestic pets were not associated with either seroprevalence or antibody titer.

**Figure 3.**
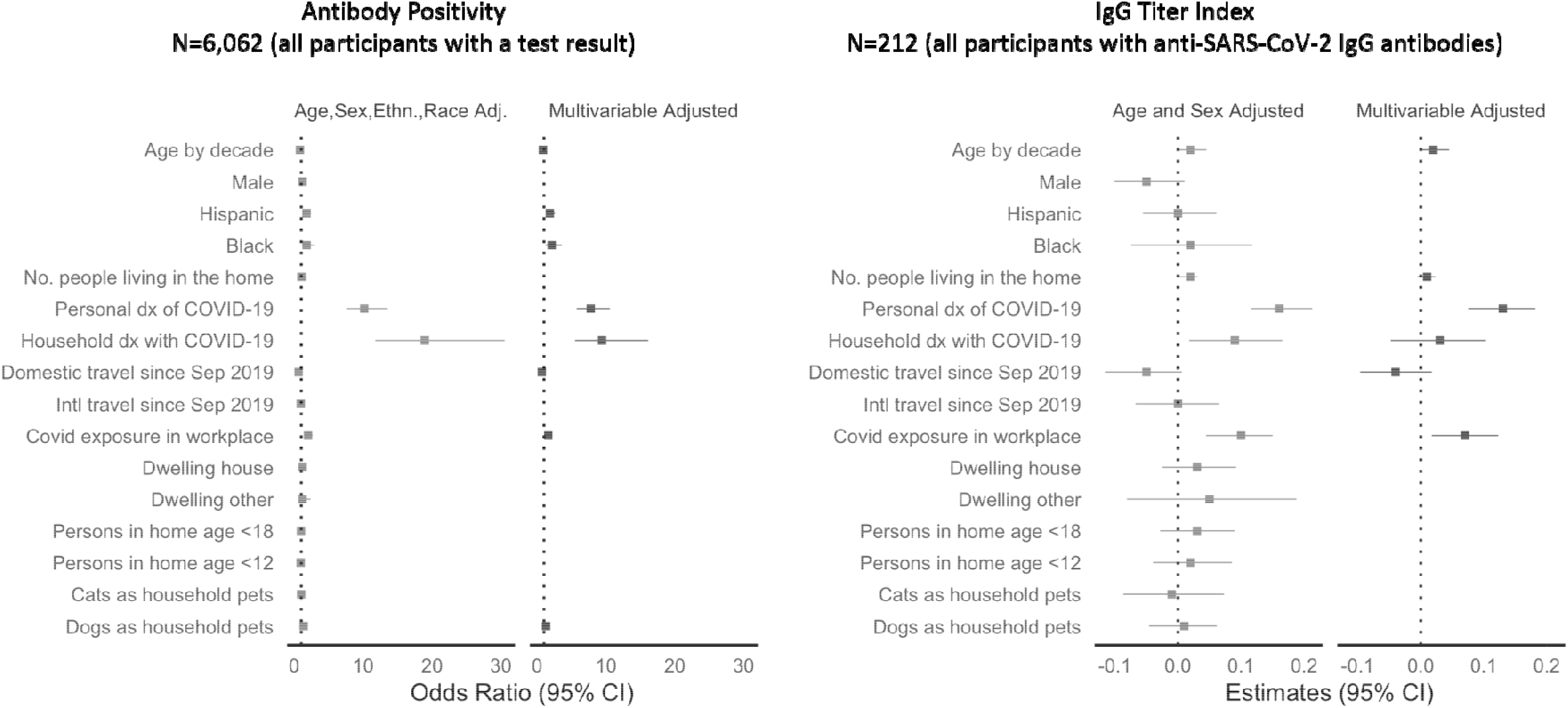
Potential COVID Illness Exposure Related Factors Associated with SARS-CoV-2.

In multivariable-adjusted analyses of Covid-19 response variables (**Figure 4** and **Supplemental Table 6**), the strongest self-reported symptom associated with greater odds of seropositive status was anosmia (11.53 [7.51, 17.70], P<0.001). Other symptoms associated with the presence of antibodies included dry cough, loss of appetite, and myalgias. Notably, the symptoms associated with lower odds of seropositive status included sore throat and rhinorrhea. Dyspnea was significantly associated with higher titer levels in seropositive individuals (beta 0.13 [SE 0.04], P=0.001).

**Figure 4.**
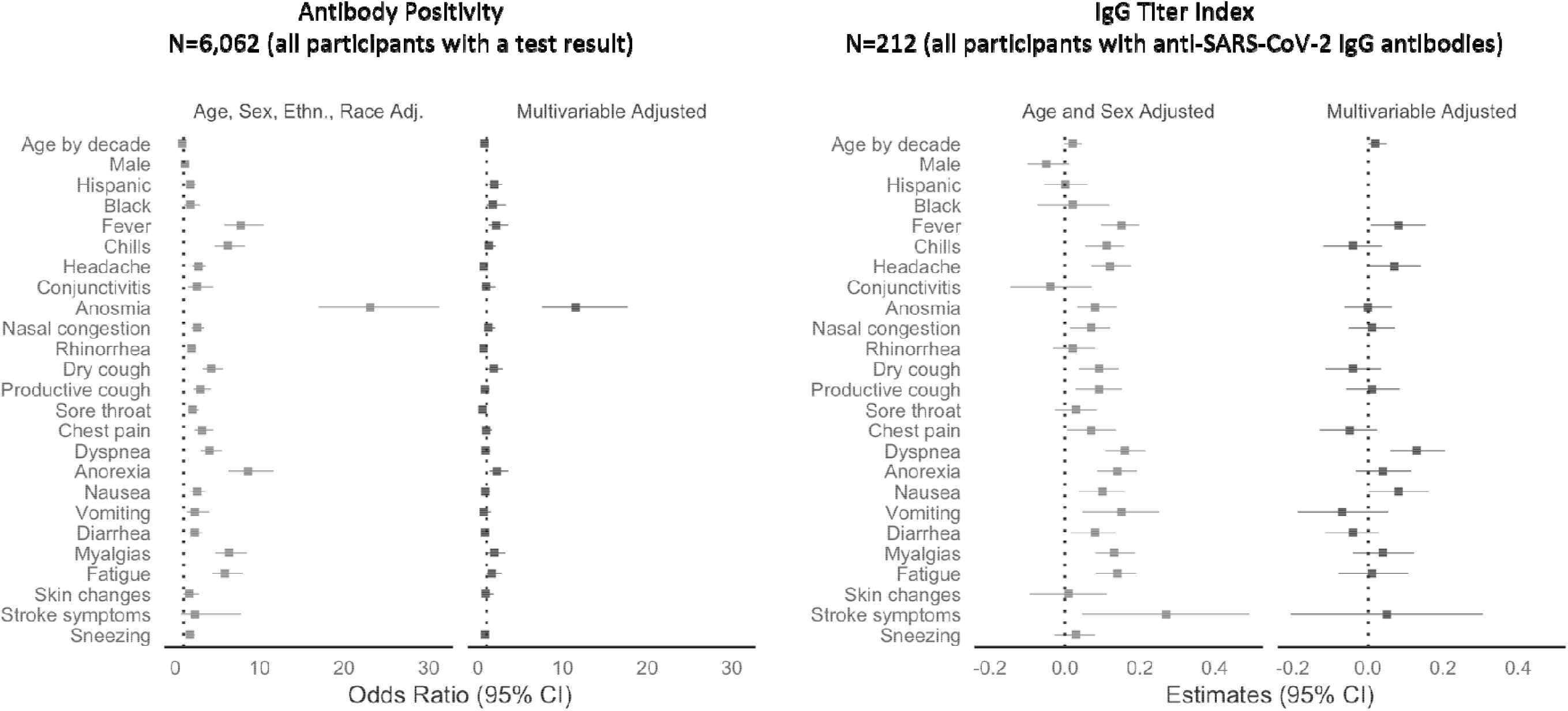
Potential COVID Illness Response Factors Associated with SARS-CoV-2 Seroprevalence.

Significantly predictive pre-existing characteristics, exposures and symptoms from the prior models were subsequently analyzed together. In multivariable analysis, all included predictors, except for dry cough, myalgias and fatigue remained significantly associated with the presence of antibodies. Predictors which remained significantly associated with higher antibody levels included hypertension (beta 0.09 [SE 0.04], P=0.031), prior Covid-19 diagnosis (beta 0.09 [SE 0.03], P=0.002), working in a Covid unit (beta 0.07 [SE 0.03], P=0.008), and dyspnea (beta 0.07 [SE 0.03], P=0.015) (**Figure 5** and **Supplemental Table 7**).

**Figure 5.**
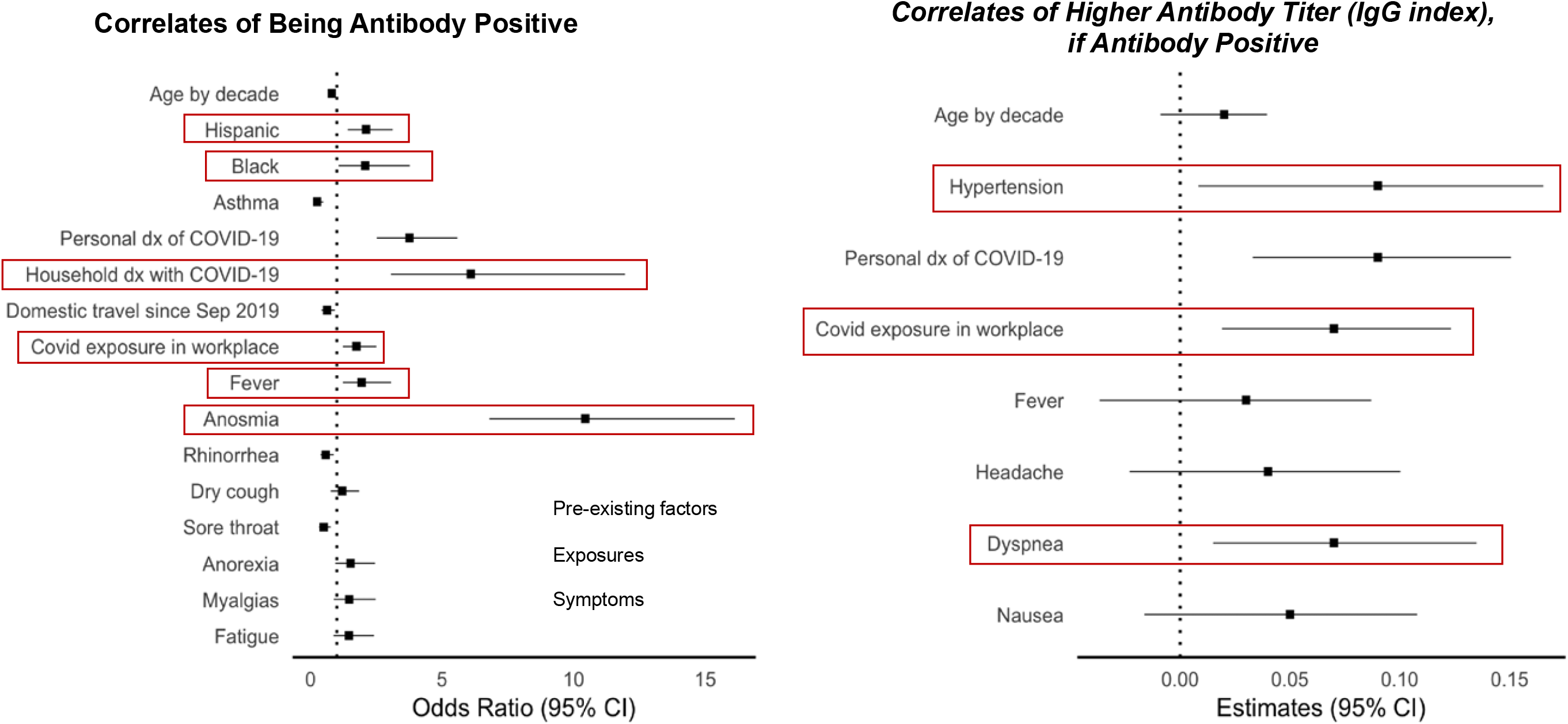
Factors Associated with SARS-CoV-2.

In a single combined model including significant pre-existing, exposure, and symptom factors, the main findings were unchanged from those in the primary analyses (**Supplemental Figure 2**).

## DISCUSSION

In a large diverse healthcare employee cohort of over 6,000 adults in Los Angeles, we observed a seroprevalence rate of 4.1%, which when accounting for published test characteristics, may range from 3.1% to 5.7%. Seroprevalence varied across demographic, clinical, exposure and symptom based characteristics. Specifically, factors significantly associated with presence of IgG antibodies included younger age, Hispanic ethnicity, and African-American race, as were exposure related factors including the presence of either a personal or household member having a prior medical diagnosis of Covid-19. Among self-reported symptoms, anosmia was most strongly associated with the presence of antibodies, with positive associations also noted for fever, dry cough, anorexia, and myalgias. The size and diversity of this study population, combined with robust survey and modeling techniques, provide a more vibrant picture of the population at highest risk for Covid-19 infection, risks of various potential exposures and symptoms that should alter patients to potential illness.

Most prior seroprevalence studieshave focused on cohorts that included healthcare workers predominantly involved in direct or indirect patient care, persons living within a circumscribed region with high viral exposure rates, or larger geographic areas from which motivated individuals could voluntarily enroll into community screening programs.^14,15^ Given that completely unbiased population-scale sampling for seroprevalence studies remains a logistical challenge, we used a sampling approach that involved open enrollment and convenient access to testing facilities made available to all employees working across multiple sites of a large healthcare system; this approach was intended to broadly capture individuals with both patient-related exposures and community-related exposures, while also representative of a relatively wide geographic area in and around Los Angeles County. Although limited to persons who are generally healthy and able to be employed, our study cohort included individuals representing a diversity of demographic characteristics including ethnicity and race – leading to findings that reflect the disparities that have been persistently observed and reported for Covid-19 infection rates in our local communities.

Consistent with findings from studies in healthcare workers, seroprevalence patterns in our cohort indicate exposure from not only the work environment but also from the home environment and likely unmeasured community-based factors.^16^ It has been well reported that minority populations, particularly African Americans and Hispanics, have been disproportionately effected by the Covid-19 panedmic.^17-19^ Our study is consistent with these prior findings, but demonstrates that such differences exist even when all participants work not just in the same field, but for the same organization. Such a finding may indicate that community and non-work related environmental factors are likely playing a significant role in the spread of Covid-19 among certain minority populations. Even after controlling for a medical diagnosis of Covid-19, African American race and Hispanic ethnicity remained risk factors for antibody positivity. The persistence of thse racial and ethnic disparities may represent structural barriers to care or societally mediated risk. Geographic clustering by race and ethnicity in housing, shopping and social gatherings may be one such factor, while socioeconomic status and ability to self-isolate outside of work likely also contribute.^20-22^

No self-reported pre-existing medical conditions were significantly associated with antibody positivity, indicating that infection itself is agnostic to baseline health. In fact, asthma was negatively associated with the presence of antibodies, or at least antibody levels above the current threshold we use for positivity. While reactive airway disease is unlikely a protective factor against Covid-19, participants with such conditions may be more likely to deligently follow social distancing guidelines and practice better adherence to hand hygiene and use of personal protective equipment. Hypertension was the only medical condition associated with higher SARS-CoV-2 antibody levels. It remains unclear as to what physiologic mechanism may contribute to this finding, however, unmeasured confounding variables, such as medications or renal disease may function as mediating factors. Further studies will be needed to both verify and elucidate this finding.

Also concordant with prior studies, we found that anosmia was the single strongest symptom associated with SARS-CoV-2 IgG antibody presence.^23-25^ Interestingly, neither dyspnea nor diarrhea, two commonly cited symptoms, demonstrated a significant association in multivariable analysis.^26,27^ This is likely related to the non-specific nature of these symptoms, which are common to multiple viral and non-viral etiologies. Importantly, dyspnea was associated with a higher antibody level among those with anti-SARS-CoV-2 antibodies, suggesting that dyspnea related to Covid-19 may drive a more robust humoral immune response, potentially related to more severe infection. These findings are concordant with the known phenomenon of proportionate adaptive immune response to higher doses of antigenic stress.^28^ The extent to which the generation of measurably higher antibody levels could confer immunity to a larger degree or for a longer duration of time remains unknown. Interestingly, prior studies have demonstrated lower antibody levels among exposed, asymptomatic individuals, a phenomena which may be attributable to a highly efficient cell mediated immune response.^29^ It has be suggested that higher T-cell levels, whether virus specific or otherwise, may play a role in this finding, however, further research is required.^30,31^

Further expanding from prior studies, we investigated and observed several factors that appeared notably unassociated with seroprevalence. In particular, we found that recent travel, type of home, and number of people living in the home were not associated with an antibody-based measure of SARS-CoV-2 exposure. The presence of antibodies was also not related to youth or children in the home, or to having domestic pets such as cats or dogs. Although far from definitive, these results suggest that these factors do not play an important role in mediating potentially meaningful viral exposure in the communities represented by our study cohort.

Several limitations of this study merit consideration. Of the employees actively employed at our multi-site institution, only a proportion of all eligible participants enrolled; nonetheless, the sample size of the cohort was large, diverse, and representative of the source sample.^7^ Our seroprevalence estimates were based on using a validated assay of only IgG antibodies; assays of IgM antibodies may offer complementary information in future studies. Data collected on medical history, exposures, and symptoms were all self-reported, similar to approaches used in prior studies. We were unable to completely verify prior Covid-19 illness using viral test results in part given lack of universally available testing for all individuals, particularly those with minimal to no symptoms.

In conclusion, in a highly diverse population of healthcare workers, demographic factors associated with Covid-19 antibody positivity indicate potential factors outside of the workplace associated with SARS-CoV-2 exposure, although these do not appear related to the number of people or to the presence of children in the home. Further, while for dyspnea may be a marker of more severe disease among those with Covid-19, it’s presence alone does not indicate infection.

## Data Availability

The data that support the findings of this study are available from Cedars-Sinai Medical Center, upon reasonable request. The data are not publicly available due to the contents including information that could compromise research participant privacy/consent.

## ACKNOWLEDGEMENTS

We are grateful to all the front-line healthcare workers in our healthcare system who continue to be dedicated to delivering the highest quality care for all patients.

## SUPPLEMENTAL MATERIAL

**Supplemental Figure 1.**
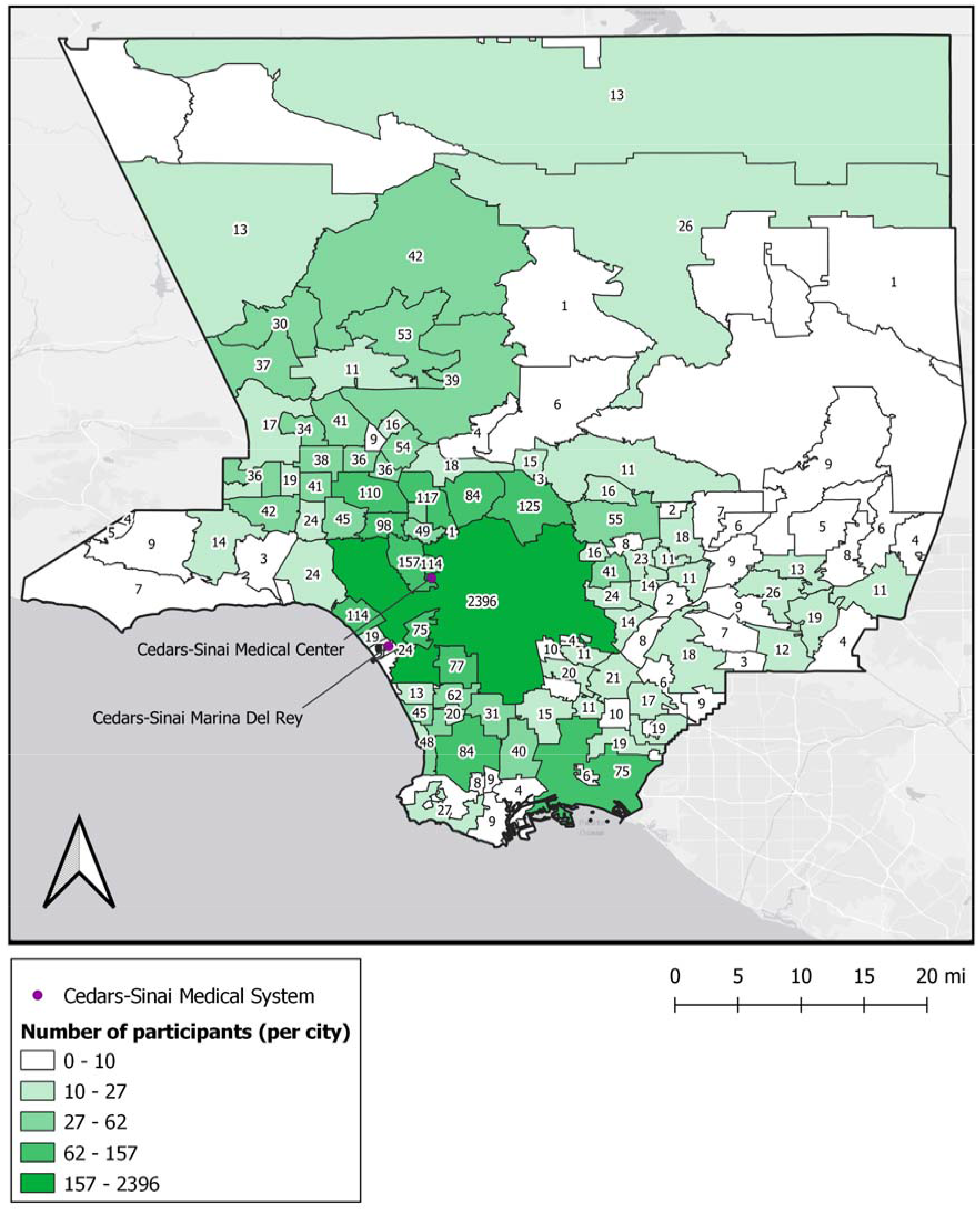

**Supplemental Table 1.**
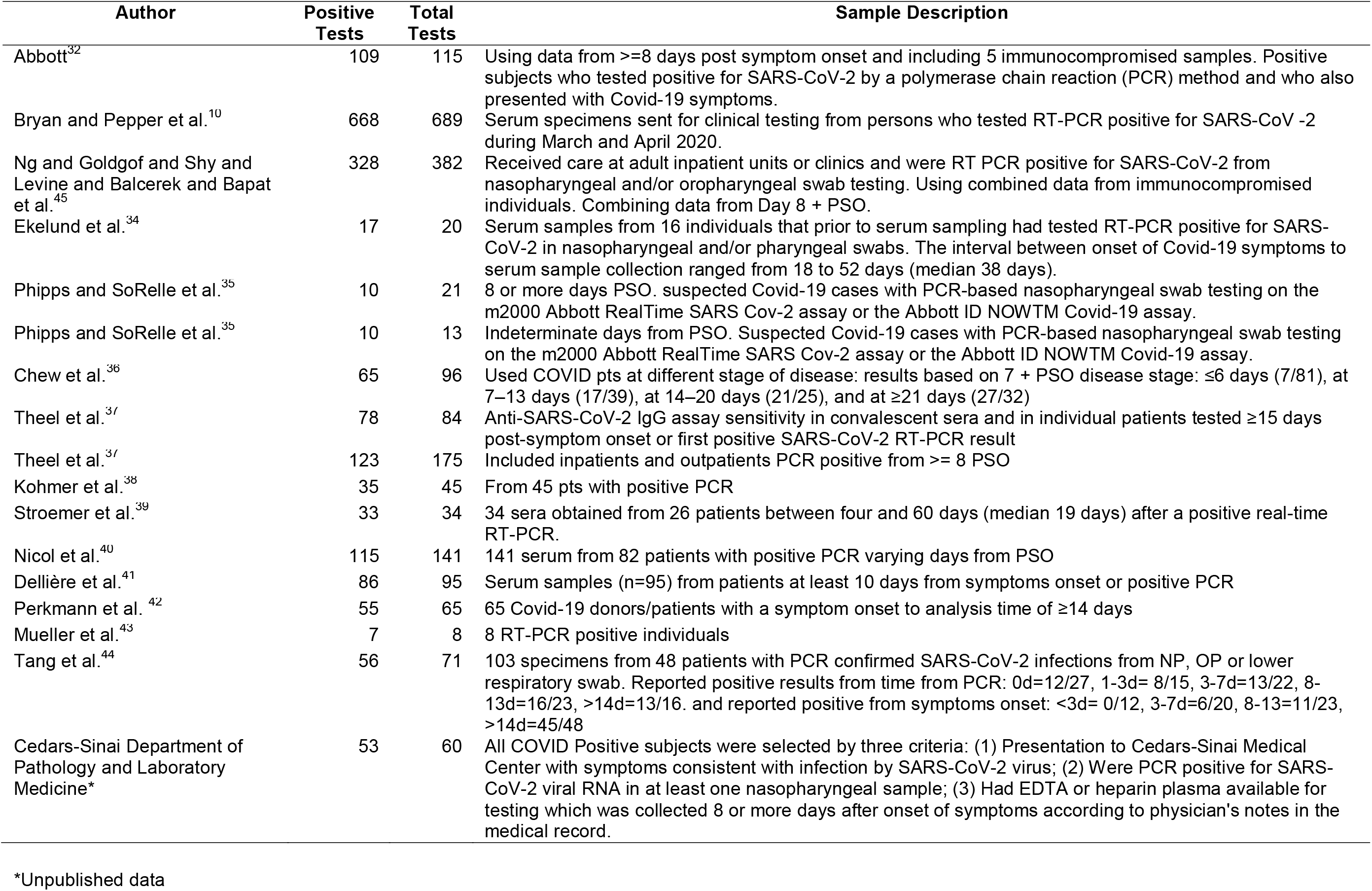
Prior Studies Reporting Sensitivity for the Abbott Architect SARS-CoV-2 IgG Assay^10,32-44^

**Supplemental Table 2.**
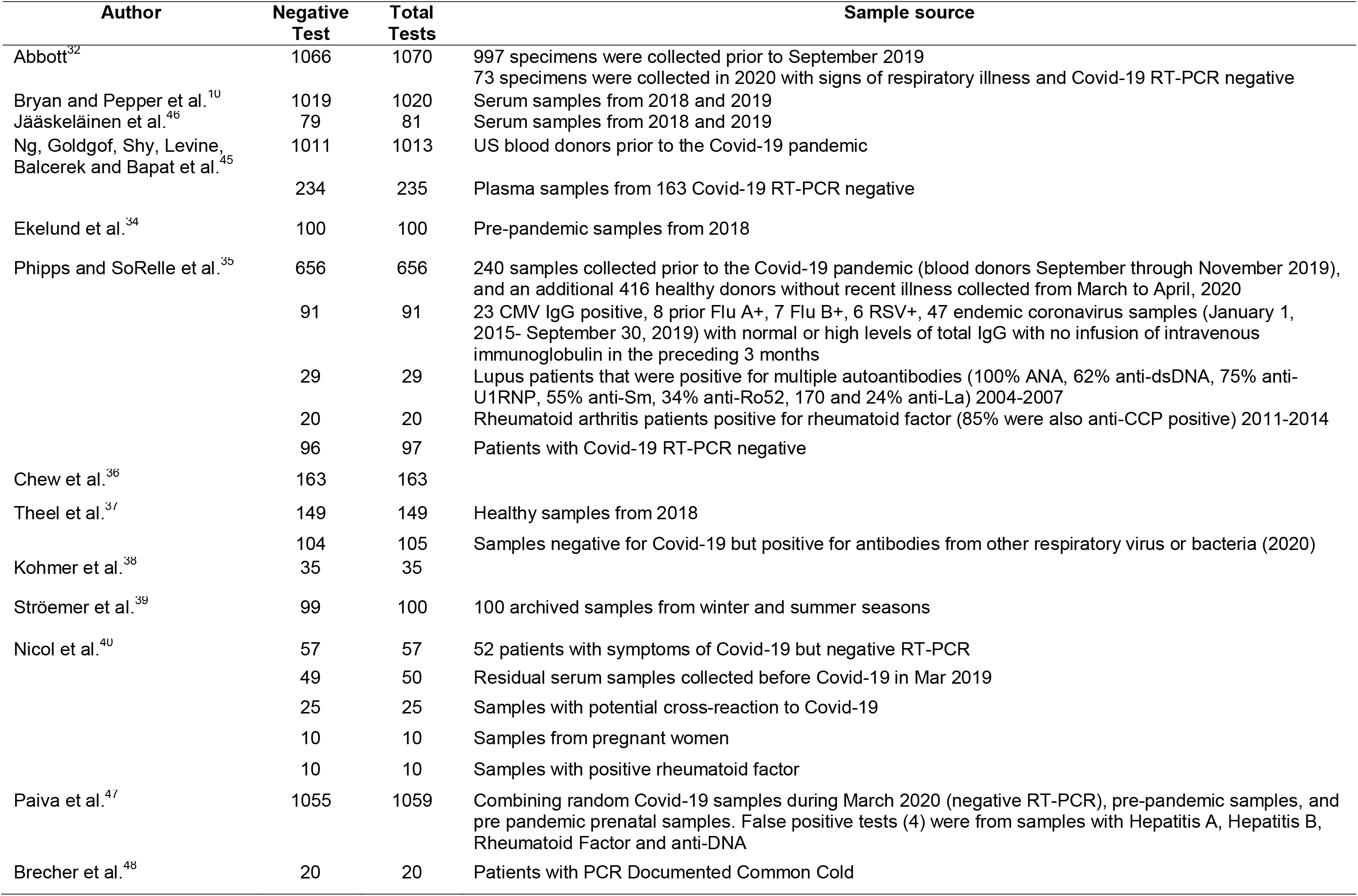
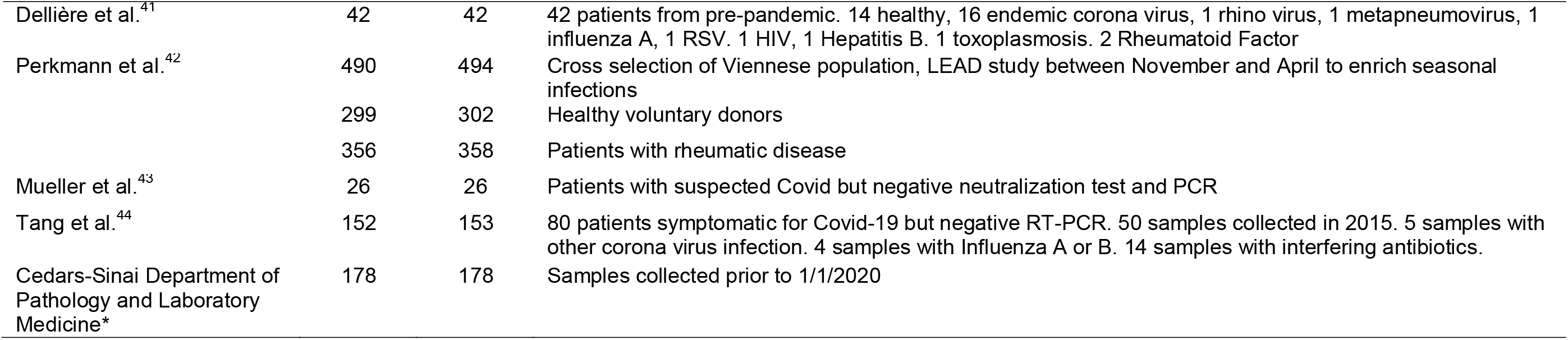
Prior Studies Reporting Specificity for the Abbott Architect SARS-CoV-2 IgG Assay

| Author | Negative Test | Total Tests | Sample source |
| --- | --- | --- | --- |
| Abbott<sup>32</sup> | 1066 | 1070 | 997 specimens were collected prior to September 2019<br>73 specimens were collected in 2020 with signs of respiratory illness and Covid-19 RT-PCR negative |
| Bryan and Pepper et al.<sup>10</sup> | 1019 | 1020 | Serum samples from 2018 and 2019 |
| Jääskeläinen et al.<sup>46</sup> | 79 | 81 | Serum samples from 2018 and 2019 |
| Ng, Goldgof, Shy, Levine, Balcerek and Bapat et al.<sup>45</sup> | 1011 | 1013 | US blood donors prior to the Covid-19 pandemic |
|  | 234 | 235 | Plasma samples from 163 Covid-19 RT-PCR negative |
| Ekelund et al.<sup>34</sup> | 100 | 100 | Pre-pandemic samples from 2018 |
| Phipps and SoRelle et al.<sup>35</sup> | 656 | 656 | 240 samples collected prior to the Covid-19 pandemic (blood donors September through November 2019), and an additional 416 healthy donors without recent illness collected from March to April, 2020 |
|  | 91 | 91 | 23 CMV IgG positive, 8 prior Flu A+, 7 Flu B+, 6 RSV+, 47 endemic coronavirus samples (January 1, 2015- September 30, 2019) with normal or high levels of total IgG with no infusion of intravenous immunoglobulin in the preceding 3 months |
|  | 29 | 29 | Lupus patients that were positive for multiple autoantibodies (100% ANA, 62% anti-dsDNA, 75% anti-U1RNP, 55% anti-Sm, 34% anti-Ro52, 170 and 24% anti-La) 2004-2007 |
|  | 20 | 20 | Rheumatoid arthritis patients positive for rheumatoid factor (85% were also anti-CCP positive) 2011-2014 |
|  | 96 | 97 | Patients with Covid-19 RT-PCR negative |
| Chew et al.<sup>36</sup> | 163 | 163 |  |
| Theel et al.<sup>37</sup> | 149 | 149 | Healthy samples from 2018 |
|  | 104 | 105 | Samples negative for Covid-19 but positive for antibodies from other respiratory virus or bacteria (2020) |
| Kohmer et al.<sup>38</sup> | 35 | 35 |  |
| Ströemer et al.<sup>39</sup> | 99 | 100 | 100 archived samples from winter and summer seasons |
| Nicol et al.<sup>40</sup> | 57 | 57 | 52 patients with symptoms of Covid-19 but negative RT-PCR |
|  | 49 | 50 | Residual serum samples collected before Covid-19 in Mar 2019 |
|  | 25 | 25 | Samples with potential cross-reaction to Covid-19 |
|  | 10 | 10 | Samples from pregnant women |
|  | 10 | 10 | Samples with positive rheumatoid factor |
| Paiva et al.<sup>47</sup> | 1055 | 1059 | Combining random Covid-19 samples during March 2020 (negative RT-PCR), pre-pandemic samples, and pre pandemic prenatal samples. False positive tests (4) were from samples with Hepatitis A, Hepatitis B, Rheumatoid Factor and anti-DNA |
| Brecher et al.<sup>48</sup> | 20 | 20 | Patients with PCR Documented Common Cold |
| Dellière et al.<sup>41</sup> | 42 | 42 | 42 patients from pre-pandemic. 14 healthy, 16 endemic corona virus, 1 rhino virus, 1 metapneumovirus, 1 influenza A, 1 RSV. 1 HIV, 1 Hepatitis B. 1 toxoplasmosis. 2 Rheumatoid Factor |
| Perkmann et al.<sup>42</sup> | 490 | 494 | Cross selection of Viennese population, LEAD study between November and April to enrich seasonal infections |
|  | 299 | 302 | Healthy voluntary donors |
|  | 356 | 358 | Patients with rheumatic disease |
| Mueller et al.<sup>43</sup> | 26 | 26 | Patients with suspected Covid but negative neutralization test and PCR |
| Tang et al.<sup>44</sup> | 152 | 153 | 80 patients symptomatic for Covid-19 but negative RT-PCR. 50 samples collected in 2015. 5 samples with other corona virus infection. 4 samples with Influenza A or B. 14 samples with interfering antibiotics. |
| Cedars-Sinai Department of Pathology and Laboratory Medicine* | 178 | 178 | Samples collected prior to 1/1/2020 |

**Supplemental Table 3.** Prevalence of Measurable SARS-CoV-2 IgG Antibody in the Study Sample

|  | Mean (95% CI) |
| --- | --- |
| **Overall** | **4.1 (3.1, 5.7)** |
| Sex: Female | 3.9 (3.0, 5.6) |
| Sex: Male | 4.3 (3.1, 6.3) |
| Age: <25 | 4.5 (2.4, 7.7) |
| Age: 25-29 | 5.1 (3.4, 7.7) |
| Age: 30-34 | 5.1 (3.5, 7.5) |
| Age: 35-39 | 3.6 (2.3, 5.3) |
| Age: 40-44 | 4 (2.6, 6.1) |
| Age: 45-49 | 3.2 (1.8, 5.1) |
| Age: 50-54 | 3.7 (2.1, 5.7) |
| Age: 55-59 | 3.5 (1.9, 5.6) |
| Age: 60-64 | 3.8 (2.2, 6.0) |
| Age: >65 | 3.1 (1.5, 5.1) |
| Race Eth.: Asian | 3.4 (2.4, 5.0) |
| Race Eth.: Black | 4.8 (2.8, 8.0) |
| Race Eth.: Hispanic / Latino | 5.7 (3.9, 8.3) |
| Race Eth.: Other | 3.4 (1.8, 5.4) |
| Race Eth.: White | 3.1 (2.1, 4.5) |

**Supplemental Table 4.** Pre-Existing Factors Associated with SARS-CoV-2 Seroprevalence

| Predictors | Outcome: Antibody Positive<br>N=6,062 (everybody with a test result) |  |  |  | Outcome: IgG index (divided by 10)<br>N=212 (everybody with a test result) |  |  |  |
| --- | --- | --- | --- | --- | --- | --- | --- | --- |
|  | Model 1 |  | Model 2 |  | Model 3 |  | Model 4 |  |
|  | OR (95% CI) | P | OR (95% CI) | P | Est (SE) | P | Est (SE) | P |
| Age (per decade) | 0.8 (0.7, 0.91) | 0.001 | 0.81 (0.71, 0.92) | 0.001 | 0.02 (0.01) | 0.07 | 0.01 (0.01) | 0.43 |
| Male Sex | 1.19 (0.89, 1.59) | 0.24 |  |  | -0.05 (0.03) | 0.11 |  |  |
| Hispanic Ethnicity | 1.76 (1.28, 2.40) | <0.001 | 1.8 (1.31, 2.46) | <0.001 | 0 (0.03) | 0.93 |  |  |
| African American Race | 1.77 (1.07, 2.93) | 0.027 | 1.72 (1.03, 2.89) | 0.04 | 0.02 (0.05) | 0.66 |  |  |
| Smoking | 0.83 (0.26, 2.66) | 0.76 |  |  | -0.01 (0.11) | 0.91 |  |  |
| Vaping | 1.12 (0.4, 3.12) | 0.82 |  |  | -0.08 (0.1) | 0.45 |  |  |
| Asthma | 0.48 (0.28, 0.83) | 0.009 | 0.48 (0.28, 0.8) | 0.009 | 0.02 (0.05) | 0.71 |  |  |
| Immune Disorder | 0.5 (0.18, 1.35) | 0.17 |  |  | -0.07 (0.1) | 0.49 |  |  |
| Cancer | 0.54 (0.17, 1.72) | 0.29 |  |  | 0.01 (0.12) | 0.92 |  |  |
| Cardiovascular Disease | 0.49 (0.12, 2.02) | 0.33 |  |  | 0.06 (0.14) | 0.65 |  |  |
| Chronic Obstructive Pulmonary Disease | 0 (0, Inf) | 0.97 |  |  |  |  |  |  |
| Diabetes Mellitus | 0.66 (0.32, 1.37) | 0.26 |  |  | 0.07 (0.07) | 0.31 |  |  |
| Hypertension | 0.9 (0.58, 1.41) | 0.64 |  |  | 0.11 (0.04) | 0.013 | 0.11 (0.04) | 0.011 |

Model 1 is adjusted for age, sex, ethnicity, race.
Model 2 is adjusted for anything that was significant in Model 1 to a P<0.10.
Model 3 is for age, sex
Model 4 is adjusted for anything that was significant in Model 3 to a P<0.10.

**Supplemental Table 5.**
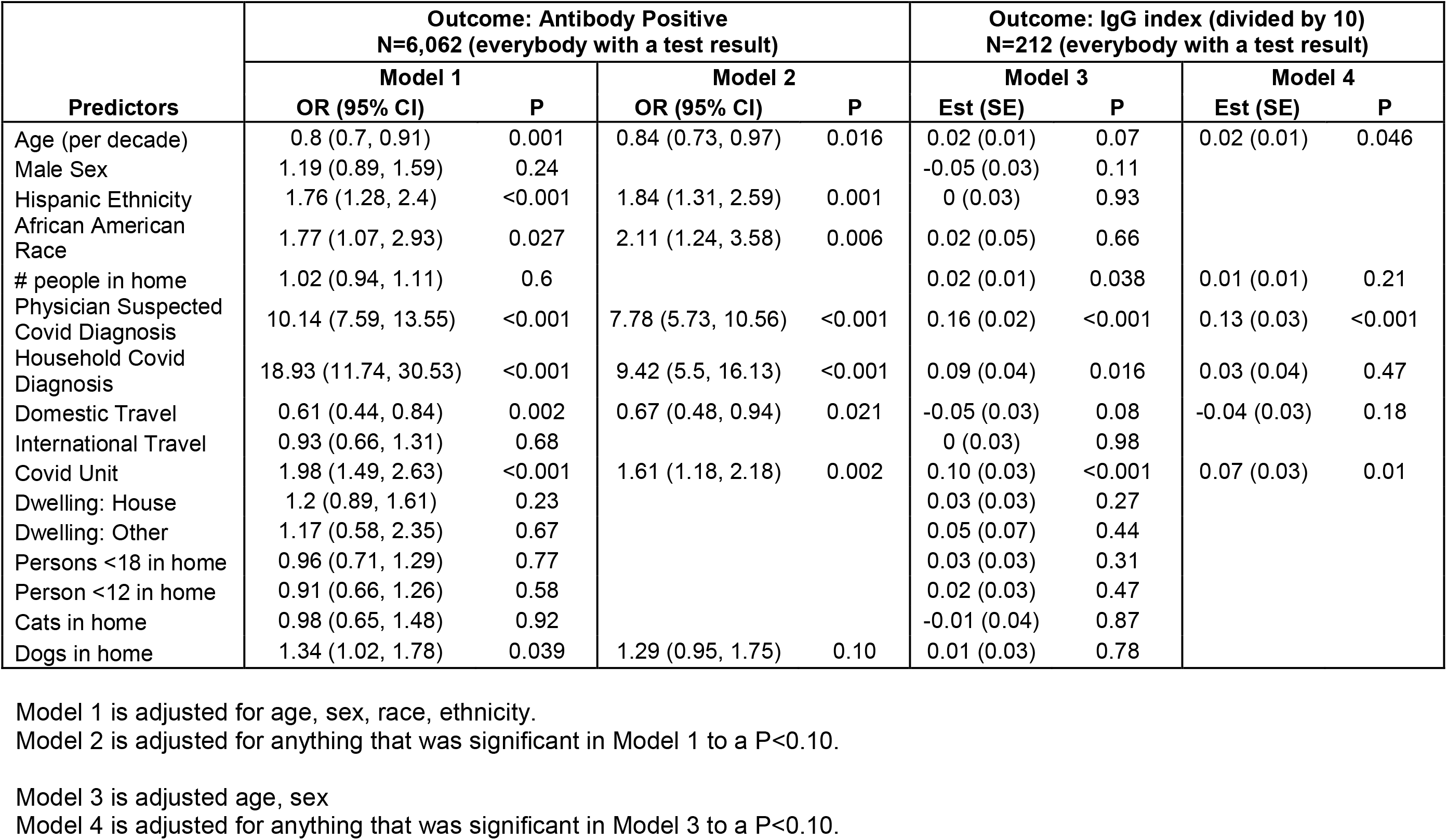
Potential COVID Illness Exposure Related Factors Associated with SARS-CoV-2 Seroprevalence

| Predictors | Outcome: Antibody Positive<br>N=6,062 (everybody with a test result) |  |  |  | Outcome: IgG index (divided by 10)<br>N=212 (everybody with a test result) |  |  |  |
| --- | --- | --- | --- | --- | --- | --- | --- | --- |
|  | Model 1 |  | Model 2 |  | Model 3 |  | Model 4 |  |
|  | OR (95% CI) | P | OR (95% CI) | P | Est (SE) | P | Est (SE) | P |
| Age (per decade) | 0.8 (0.7, 0.91) | 0.001 | 0.84 (0.73, 0.97) | 0.016 | 0.02 (0.01) | 0.07 | 0.02 (0.01) | 0.046 |
| Male Sex | 1.19 (0.89, 1.59) | 0.24 |  |  | -0.05 (0.03) | 0.11 |  |  |
| Hispanic Ethnicity | 1.76 (1.28, 2.4) | <0.001 | 1.84 (1.31, 2.59) | 0.001 | 0 (0.03) | 0.93 |  |  |
| African American Race | 1.77 (1.07, 2.93) | 0.027 | 2.11 (1.24, 3.58) | 0.006 | 0.02 (0.05) | 0.66 |  |  |
| # people in home | 1.02 (0.94, 1.11) | 0.6 |  |  | 0.02 (0.01) | 0.038 | 0.01 (0.01) | 0.21 |
| Physician Suspected Covid Diagnosis | 10.14 (7.59, 13.55) | <0.001 | 7.78 (5.73, 10.56) | <0.001 | 0.16 (0.02) | <0.001 | 0.13 (0.03) | <0.001 |
| Household Covid Diagnosis | 18.93 (11.74, 30.53) | <0.001 | 9.42 (5.5, 16.13) | <0.001 | 0.09 (0.04) | 0.016 | 0.03 (0.04) | 0.47 |
| Domestic Travel | 0.61 (0.44, 0.84) | 0.002 | 0.67 (0.48, 0.94) | 0.021 | -0.05 (0.03) | 0.08 | -0.04 (0.03) | 0.18 |
| International Travel | 0.93 (0.66, 1.31) | 0.68 |  |  | 0 (0.03) | 0.98 |  |  |
| Covid Unit | 1.98 (1.49, 2.63) | <0.001 | 1.61 (1.18, 2.18) | 0.002 | 0.10 (0.03) | <0.001 | 0.07 (0.03) | 0.01 |
| Dwelling: House | 1.2 (0.89, 1.61) | 0.23 |  |  | 0.03 (0.03) | 0.27 |  |  |
| Dwelling: Other | 1.17 (0.58, 2.35) | 0.67 |  |  | 0.05 (0.07) | 0.44 |  |  |
| Persons <18 in home | 0.96 (0.71, 1.29) | 0.77 |  |  | 0.03 (0.03) | 0.31 |  |  |
| Person <12 in home | 0.91 (0.66, 1.26) | 0.58 |  |  | 0.02 (0.03) | 0.47 |  |  |
| Cats in home | 0.98 (0.65, 1.48) | 0.92 |  |  | -0.01 (0.04) | 0.87 |  |  |
| Dogs in home | 1.34 (1.02, 1.78) | 0.039 | 1.29 (0.95, 1.75) | 0.10 | 0.01 (0.03) | 0.78 |  |  |

Model 1 is adjusted for age, sex, race, ethnicity.
Model 2 is adjusted for anything that was significant in Model 1 to a P<0.10.
Model 3 is adjusted age, sex
Model 4 is adjusted for anything that was significant in Model 3 to a P<0.10.

**Supplemental Table 6.**
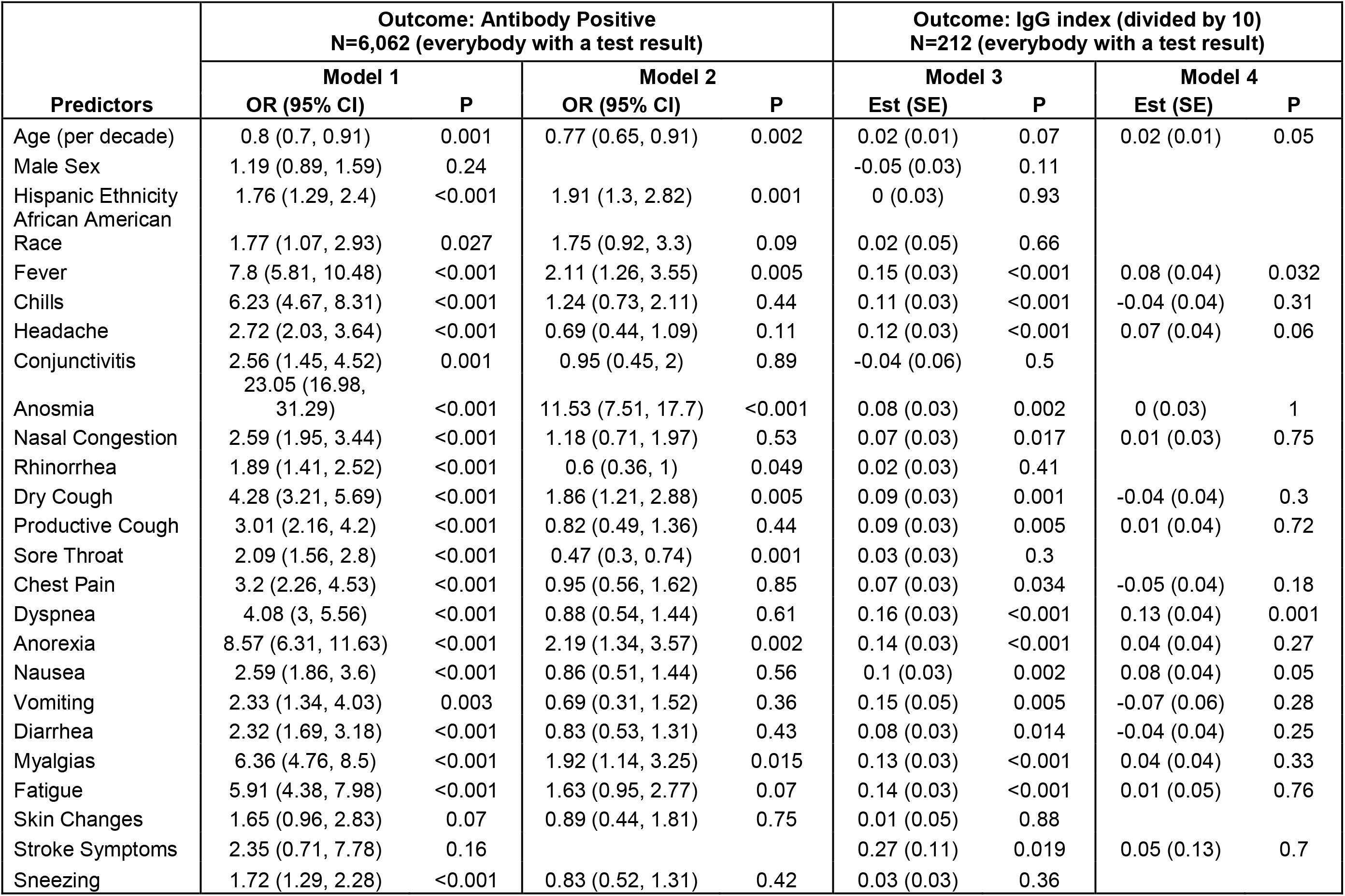
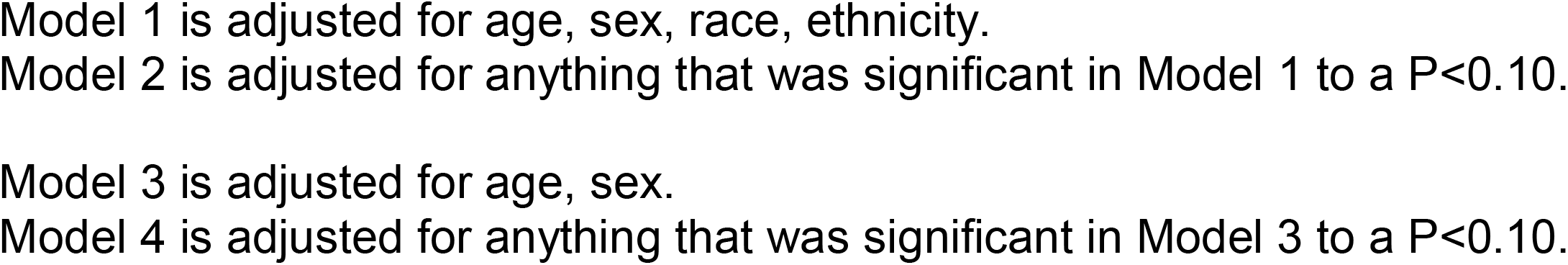
Potential COVID Illness Response Factors Associated with SARS-CoV-2 Seroprevalence

**Supplemental Table 7.** Factors Associated with SARS-CoV-2

| Predictors | Outcome: Antibody Positive<br>N=6,062 (everybody with a test result) |  | Outcome: IgG index (divided by 10)<br>N=212 (everybody with a test result) |  |
| --- | --- | --- | --- | --- |
|  | OR (95% CI) | P | Est (SE) | P |
| Age (per decade) | 0.81 (0.69, 0.96) | 0.017 | 0.02 (0.01) | 0.22 |
| Hispanic Ethnicity | 2.11 (1.43, 3.13) | <0.001 |  |  |
| African American Race | 2.08 (1.12, 3.88) | 0.021 |  |  |
| Asthma | 0.25 (0.13, 0.5) | <0.001 |  |  |
| Hypertension |  |  | 0.09 (0.04) | 0.031 |
| Physician Suspected Covid Diagnosis | 3.76 (2.52, 5.59) | <0.001 | 0.09 (0.03) | 0.002 |
| Household Covid Diagnosis | 6.09 (3.08, 12.06) | <0.001 |  |  |
| Domestic Travel | 0.63 (0.42, 0.92) | 0.019 |  |  |
| Covid Unit | 1.75 (1.23, 2.5) | 0.002 | 0.07 (0.03) | 0.008 |
| Fever | 1.94 (1.23, 3.07) | 0.004 | 0.03 (0.03) | 0.42 |
| Headache |  |  | 0.04 (0.03) | 0.22 |
| Anosmia | 10.44 (6.78, 16.07) | <0.001 |  |  |
| Rhinorrhea | 0.58 (0.38, 0.89) | 0.012 |  |  |
| Dry Cough | 1.2 (0.77, 1.86) | 0.42 |  |  |
| Sore Throat | 0.5 (0.32, 0.77) | 0.002 |  |  |
| Dyspnea |  |  | 0.07 (0.03) | 0.015 |
| Anorexia | 1.52 (0.94, 2.46) | 0.09 |  |  |
| Nausea |  |  | 0.05 (0.03) | 0.15 |
| Myalgias | 1.47 (0.88, 2.48) | 0.14 |  |  |
| Fatigue | 1.46 (0.87, 2.44) | 0.15 |  |  |

Models are adjusted for significant predictors from the primary multivariable models examining associations of existing characteristics, exposures and symptoms with antibody positivity and IgG titer index.

